# Baseline associations between exposure to metals and systolic and diastolic blood pressure among women in the Household Air Pollution Intervention Network Trial

**DOI:** 10.1101/2025.01.21.25320894

**Authors:** Patrick Karakwende, Dana Boyd Barr, William Checkley, Thomas Clasen, Amy Lovvorn, Carmen Lucía Contreras, Anaite A. Diaz, Ephrem Dusabimana, Lisa De Las Fuentes, Shirin Jabbarzadeh, Michael Johnson, Egide Kalisa, Miles Kirby, John P. McCracken, Florien Ndagijimana, Adolphe Ndikubwimana, Theoneste Ntakirutimana, Jean de Dieu Ntivuguruzwa, L Jennifer, Ajay Pillarisetti Peel, Victor G. Davila-Roman, Ghislaine Rosa, Lance A. Waller, Jiantong Wang, Lisa Thompson, Maggie L. Clark, Bonnie N. Young the HAPIN investigators, the HAPIN investigators

## Abstract

Lead (Pb) and cadmium (Cd) are metals that occur naturally in the environment and are present in biomass fuels, such as wood. When these fuels are burned, they can release Pb and Cd into the air, leading to exposure through inhalation. Studies of exposure to metals and health outcomes suggest harmful impacts, including cardiovascular diseases. We assessed baseline associations between Pb and Cd concentrations in dried blood spots with systolic and diastolic blood pressure (SBP, DBP) among women in the Household Air Pollution Intervention Network (HAPIN) trial. We analyzed data from three of the four HAPIN randomized controlled trial sites (Guatemala, Peru, and Rwanda), focusing on women aged 40 to 79 years living in households reliant on biomass cooking. Dried blood spots were collected, processed, and analyzed for Pb and Cd exposure; SBP and DBP were measured following international guidelines. Demographic, socioeconomic, and dietary variables were collected via standardized questionnaires administered by local field staff. Statistical analyses included multivariable linear regression to examine associations between Pb and Cd, separately, and BP, adjusting for covariates informed by a Directed Acyclic Graph. Additional analyses assessed effect modification by age and research site. There was regional variation in BP levels among women, with median SBP and DBP values higher in Rwanda (116.3 mmHg, 73.0 mmHg) and Guatemala (113.3 mmHg, 68.3 mmHg) compared to Peru (106.0 mmHg, 63.3 mmHg). Pb exposure showed positive associations with both SBP and DBP. For each log-unit increase in Pb concentration, we observed increases of 2.36 mmHg SBP (95% CI 0.51, 4.20) and 1.42 mmHg DBP (95% CI 0.16, 2.67). Cd was not associated with SBP or DBP in this analysis. Pb exposure may be an important risk factor for increased SBP and DBP, markers of cardiovascular disease risk.

## 1. Introduction

Lead (Pb) and cadmium (Cd) are metals that occur naturally in the environment and are in biomass fuels, such as wood. When burned, these fuels can release Pb and Cd into the air, leading to exposure^1^. Studies on associations between exposure to these metals and health conditions suggest harmful impacts on health including cardiovascular diseases, as exposure to both Pb and Cd have been shown to be considered coronary risk factors^2^. Evidence indicates that metal levels in the body change with age due to differences in exposure sources, absorption rates, and cumulative effects^3^. Adults, particularly older adults, may experience chronic health effects due to long-term exposure and bioaccumulation^4^. Cd is considered a toxic pollutant due to its carcinogenic and mutagenic properties. Exposure to even low levels of Cd can affect the brain, kidneys, and heart and even result in death^5^. In a systematic review conducted by Bret Ericson et *al* identified major sources of lead exposure as informal lead acid battery recycling and manufacture, metal mining and processing, electronic waste, and the use of lead as a food adulterant, primarily in spices among others^6^.

There are a few suggested pathways for how Pb and Cd exposure can impact cardiovascular disease risk. Pb and Cd can lead to problems with blood coagulation^7^ and can disrupt the normal functioning of blood vessels and the heart. Pb, for example, can interfere with the regulation of blood pressure (BP) by affecting kidney function and the nervous system^8^. Exposure to these metals has been associated with an increased risk of hypertension, a major risk factor for heart attacks and strokes^9^. Chronic exposure to Pb and Cd may lead to sustained increases in BP, contributing to long-term health issues^10^. This can also result in a cycle of health decline, where hypertension exacerbates other conditions^11^. Elevated BP can serve as an early indicator of potential health impacts from environmental exposure to these heavy metals, helping in the development of preventive measures and policies^12^.

Exposure to Pb has been linked to elevated BP, primarily due to its toxic effects on the cardiovascular system. Pb interferes with calcium signaling, which is crucial for maintaining vascular tone and blood pressure regulation. It promotes oxidative stress and inflammation, leading to endothelial dysfunction and reduced nitric oxide availability, both of which impair the relaxation of blood vessels. Additionally, Pb can stimulate the sympathetic nervous system, increasing heart rate and vascular resistance. Chronic exposure may also impair kidney function, disrupting fluid and electrolyte balance, further contributing to hypertension. These combined mechanisms underscore the role of Pb exposure as a risk factor for high blood pressure^13^.

Our aim was to assess baseline associations of Pb and Cd concentrations from dried blood spots with systolic and diastolic BP (SBP, DBP) among women aged 40-79 years among 3 research sites (Guatemala, Peru, Rwanda) of the Household Air Pollution Intervention Network (HAPIN) trial. We hypothesized that SBP and DBP would be positively associated increases with higher Pb and Cd concentrations.

## 2. Materials and Methods

### 2.1. Study site and population

This paper analyzed data from the HAPIN trial from 3 of the 4 countries (Guatemala, Peru, and Rwanda, excluding India), which has been described in detail by Clasen *et al*^14^. Briefly, the HAPIN trial was a randomized controlled trial to investigate the effect of a liquefied petroleum gas (LPG) cookstove and fuel intervention on exposure to household air pollution and health. For this analysis, we assessed the association of Pb and Cd exposure on BP in women residing with pregnant women enrolled in the HAPIN trial. This analysis included non-pregnant adult female participants at baseline (before households were randomly assigned to intervention or control arms, stratified by country) in the HAPIN trial research sites (Guatemala, Peru and Rwanda). Personal exposures to metals and health outcomes were obtained from 272 adult women aged 40 to 79 years (confirmed by government-issued identification card when possible) who resided in the same households as an enrolled pregnant woman in the HAPIN trial across study sites, provided they did not meet any of the following exclusion criteria: currently smoking cigarettes or other tobacco products, or planning to permanently move out of their current household in the next 12 months. Households were included in the study if they self-reported primarily using biomass fuels for cooking^15^.

The study sites were selected because solid biomass was the primary source of fuel for a large proportion of the population. The study participants in Guatemala were characterized by the use of chimney stoves and open fires; 97% used wood fuel and cooked mainly indoors^16^. In Peru, study participants came from rural households that regularly cooked using traditional biomass (usually dung mixed with straw) burning stoves^17^. In Rwanda, 80.4% of the population relied on wood or wood-based products for cooking, which was typically done on traditional open stoves in poorly ventilated kitchens, in addition to the traditional three stone fires (62.6%) or clay brazier stove (34.6%) fueled with wood (89.9%) or charcoal (8.1%) and cooking indoors (71.9%)^18^.

### 2.2. Clinical Biomarkers and BP Measurements

Following recruiting and informed consent, qualified fieldworkers visited the families at the start of the study to do baseline questionnaires and other evaluations. Dried blood spots were collected immediately after 24-hour household air pollution exposure measurements using a GE Whatman 903 Protein Saver Card and a BD contact-activated 2.0 mm lancet. Technicians performed finger-stick punctures on participants’ non-dominant hand, cleaning the site with 70% isopropanol to ensure sample integrity. Blood was collected by gently massaging the hand to promote flow, without squeezing, and drops were allowed to fall naturally onto the filter paper. After drying at room temperature for 24-48 hours, the blood spots were stored in biospecimen bags with desiccants and humidity indicator cards at -20°C, shipped on dry ice, and analyzed at Emory University. Invalid spots were marked to guide lab processing, and valid spots were carefully selected for analysis.

At Emory University, dried blood spot samples from Guatemala, Rwanda, and Peru were analyzed using inductively coupled plasma mass spectrometry (ICP-MS) after digestion with nitric acid. A range of quality controls, including laboratory blanks and reference materials, was used to ensure accuracy. Samples were standardized for blood volume and analyzed for elements such as Pb and Cd, with strict quality control measures in place. For the potassium standardization of Pb and Cd concentrations, each metal value was multiplied by 4 mmol/mL and then divided by the individual’s specific potassium value: (metal*4)/K+.

BP measurements were recorded using an Omron HEM-907XL monitor, with three measurements taken after ensuring participants had not smoked, consumed alcohol or caffeine, or cooked with biomass in the last 30 minutes^14^.

### 2.3. Other variables

Qualified fieldworkers visited the households at the beginning of the study to conduct baseline surveys and other assessments after recruitment and informed consent. Interviews and surveys were conducted in the participant’s native language by research assistants. This baseline visit included a survey covering a range of topics like cooking behaviors, household composition, socioeconomic status (SES) and demographic information like age in years that was confirmed by national ID card, education which was self-reported and split into no formal education; primary or secondary incomplete; and secondary, college or university categories. Women self-reported exposure to secondhand tobacco (yes/no). We determined dietary diversity using the Minimum Dietary Diversity for Women (MDD-W) indicator, which is based on ten food groups (FAO FHI 360, 2016). Diet diversity was considered low if the participant had consumed less than four of the ten food groups over the previous 30 days (MDD-W < 4), medium if they had consumed four or five food groups (4 ≤ MDD-W ≤ 5), and high if they had consumed more than five (MDD-W > 5). Food insecurity was assessed by the use Food Insecurity Experience Scale, developed by the Food and Agriculture Organization of the United Nations, http://www.fao.org/3/as583e/as583e.pdf.

Material items were selected as useful indicators of SES by other HAPIN investigators to be used across all publications. SES items included color TV, radio, mobile phone, bank account, and bicycle (yes/no per item).

We measured weight and height in duplicate using seca 876 electronic scales and seca 213 stadiometer platforms (seca GmbH & Co. KG., Hamburg, Germany), respectively. If the weight or height measurements differed by more than 0.5 kg or 1 cm, respectively, we collected a third measurement and the two closest readings were averaged. We computed BMI as the ratio between weight and height-squared (kg/m2).

We determined level of physical activity using the Global Physical Activity Questionnaire (GPAQ), which collects information on both vigorous and moderate-intensity activity in three settings: at work, traveling to and from places, and in recreational activities, as well as sedentary behavior (World Health Organization, 2012). From the GPAQ data, we calculated energy consumption based on Metabolic Equivalents (METs), where one MET is defined as the energy cost of sitting quietly and is equivalent to 1 kcal/kg/hour. Since caloric consumption is an estimated four times as high when moderately active and eight times as high when vigorously active, we calculated overall energy expenditure by assigning four METs to the time spent in moderate activities and eight METs to the time spent in vigorous activities (World Health Organization, 2012). We reported this estimated physical activity as MET-minutes per day.

### 2.4. Statistical Analysis

Data were managed and analyzed using R version 4.3.3 (R Foundation for Statistical Computing) and RStudio^20^. Initially, 314 non-pregnant adult women who lived in the same household as an enrolled pregnant woman were included. For data cleaning, we removed those with missing BP values (n=6), those who reported taking high BP medicine (n=33), and those who had missing Pb and Cd values (n=3). The final dataset for analysis included 272 women with valid Pb and Cd measures.

We calculated descriptive statistics for exposure, demographic, socioeconomic status and other potential confounders. We used a Directed Acyclic Graph (DAG) to help inform which covariates to investigate and which to consider for analysis (Supplemental Figure 1). Detailed exploratory data analysis assessed frequencies and mean (SD, median, Q1-Q3, min-max) of BP outcomes (systolic and diastolic), Pb and Cd exposures, and all other covariates. We used diagnostic plots of simple linear regression models for Pb and Cd measures and BP to see if the Pb and Cd measures needed to be natural log-transformed to meet the assumptions of linear regression. We highlighted covariates that lacked any variation in responses and considered creating new categories (binning) if necessary or dropping these covariates.

We used covariates from the DAG and assessed associations with SBP and DBP and Pb and Cd in crude (i.e., unadjusted) linear regression models. We explored how these covariates were correlated with one another to assess for potentially strong correlations.

We built multivariable linear regression models with women’s SBP and DBP as separate outcomes and Pb and Cd as separate exposures. We included a priori confounders (i.e., age) and potential confounders (SES and other indicators of SES, e.g., education, household assets, dietary diversity, and food insecurity). Since BMI may be a mediator (Supplemental Figure 1), we considered models with and without BMI as an adjusted variable in sensitivity analyses.

Finally, we assessed effect modification in the adjusted linear regression models by adding an interaction term between the Pb and Cd exposure variables and age (median cutoff: <51 vs. ≥51 years) and by research site (Guatemala, Peru and Rwanda).

### 2.5. Ethical considerations

The study protocol has been reviewed and approved by institutional review boards (IRBs) or Ethics Committees at Emory University (00089799), Johns Hopkins University (00007403), Sri Ramachandra Institute of Higher Education and Research (IEC-N1/16/JUL/54/49) and the Indian Council of Medical Research – Health Ministry Screening Committee (5/8/4-30/(Env)/Indo-US/2016-NCD-I), Universidad del Valle de Guatemala (146-08-2016) and Guatemalan Ministry of Health National Ethics Committee (11-2016), Asociación Benefica PRISMA (CE2981.17), the London School of Hygiene and Tropical Medicine (11664-5) and the Rwandan National Ethics Committee (No.357/RNEC/2018), and Washington University in St. Louis (201611159). The study has been registered with ClinicalTrials.gov (Identifier NCT02944682).

## 3. Results

### 3.1. Baseline characteristics

The baseline characteristics of the participants are shown in Table 1 for those analyzed with Pb and Cd measures (n=272). The majority of metals data were from Guatemala (43%) and Peru (44%), with 13% from Rwanda. The average age of participants was 52.3 (SD 7.6) years. There was a notable prevalence of overweight and obesity (n=174, 65%) among the participants. The educational level was generally low with 77% (n=203) having no formal education or primary school incomplete (Table 1).

**Table 1:** Baseline characteristics among adult women with valid Pb and Cd measures, Guatemala, Peru, and Rwanda (N=272)

| Variable | Mean (SD); Min-Max or n (%) |
| --- | --- |
| **Research sites** |  |
| Guatemala | 117 (43%) |
| Peru | 121 (44%) |
| Rwanda | 34 (13%) |
| Age at baseline (years) | 52.3 (7.6); 40.1- 73.8 |
| Age at median cutoff, binary (years) |  |
| Aged <51 | 134 (49%) |
| Aged ≥51 | 138 (51%) |
| BMI (continuous, kg/m<sup>2</sup>) | 26.8 (4.7); 15.4- 39.2 |
| BMI, binary version |  |
| Underweight, normal | 94 (35%) |
| Overweight, obese | 174 (65%) |
| Underweight (<18.5) | 8 (3.4%) |
| Normal weight (18.5 to <25) | 86 (32%) |
| Overweight (25.0 to <30) | 109 (40.6%) |
| Color TV | 132 (49%) |
| Radio | 161 (59%) |
| Mobile phone | 257 (94%) |
| Bank account | 82 (30%) |
| Bicycle | 85 (31%) |
| METs per day of physical activity | 1086 (1030); 0- 6171 |
| Food insecurity |  |
| None | 127 (48%) |
| Mild | 95 (36%) |
| Moderate/severe | 45 (17%) |
| Dietary diversity |  |
| Low | 137 (50%) |
| Medium | 102 (38%) |
| High | 33 (12%) |
| Education 2-levels |  |
| No formal education or Primary school incomplete | 203 (77%) |
| Primary school complete, Secondary school incomplete, Secondary school complete or Vocational or Some college or university | 61 (23%) |
| Exposure to secondhand tobacco smoke | 16 (6%) |

Almost all participants had mobile phones (94%), while a half of them had radios (59%) and color tv (49%) and fewer had bicycles (31%) and a bank (30%) account. The data from this study suggested that a substantial proportion of the sample faced challenges related to both food insecurity and dietary diversity. Over half of the sample reported were food insecure with mild (36%) or moderate to severe (17%) levels. Additionally, dietary diversity was low for half the sample, indicating limited access to a variety of foods (Table 1). The results also indicate that out of the total sample population, 16 individuals (6%) reported being exposed to secondhand tobacco smoke. Participants spent a substantial amount of physical exertion, as indicated by 1,086 average METs per day (SD 1,030), although the large standard deviation and the wide range of values (from 0 to 6,171 METs per day) reveal significant variation in exercise habits among individuals.

The baseline levels of Pb and Cd as presented in Supplemental table1 and Supplemental figures 2 and 3 were; Guatemala: 117 samples, Mean = 2.02 (SD = 1.63), Median = 1.49, 25th percentile = 0.938, 75th percentile = 2.42, Min-Max = 0.08-8.72, Peru: 121 samples, Mean = 1.90 (SD = 1.90), Median = 1.19, 25th percentile = 0.880, 75th percentile = 1.99, Min-Max = 0.013-11.0 and Rwanda: 34 samples, Mean = 2.30 (SD = 2.27), Median = 1.47, 25th percentile = 0.937, 75th percentile = 2.79, Min-Max = 0.165-9.07 while for Cd; Guatemala: 117 samples, Mean = 0.996 (SD = 0.446), Median = 0.993, 25th percentile = 0.709, 75th percentile = 1.25, Min-Max = 0.116-2.23, Peru: 121 samples, Mean = 1.07 (SD = 0.462), Median = 1.05, 25th percentile = 0.775, 75th percentile = 1.42, Min-Max = 0.128-2.12 and Rwanda: 34 samples, Mean = 1.00 (SD = 0.434), Median = 1.03, 25th percentile = 0.728, 75th percentile = 1.30, Min-Max = 0.0921-1.95.

The overall mean (SD) of SBP and DBP for all three research sites was 112.2 (14.5) mmHg and 67.3 (9.6) mmHg, respectively (Table 2). The BP summary results (Table 2) and boxplots of SBP and DBP by country (Figures 2 and 3) indicated regional differences in BP among the study participants. Rwanda and Guatemala generally had higher BP levels compared to Peru.

**Table 2:** Summary of SBP and DBP among adult women with valid Pb and Cd measures at baseline visit per research site, HAPIN (Guatemala, Peru, Rwanda) N=272.

|  | N | Mean (SD) | Median | 25<sup>th</sup>, 75<sup>th</sup><br>percentiles | Min-Max |
| --- | --- | --- | --- | --- | --- |
| **Systolic BP** |  |  |  |  |  |
| **All research sites** | **272** | **112.2 (14.5)** | **109.2** | **102.6, 119.3** | **81.0-187.3** |
| Guatemala | 117 | 115.9 (16.6) | 113.3 | 104.7, 121.0 | 92.3-187.3 |
| Peru | 121 | 107.4 (11.4) | 106.0 | 100.0, 113.3 | 81.0-145.3 |
| Rwanda | 34 | 116.2 (12.03) | 116.3 | 106.2, 124.3 | 92.0-140.7 |
| **Diastolic BP** |  |  |  |  |  |
| **All research sites** | **272** | **67.3 (9.6)** | **66.2** | **61.0, 73.0** | **49.0-106.3** |
| Guatemala | 117 | 69.2 (10.0) | 68.3 | 62.0, 75.0 | 49.0-106.3 |
| Peru | 121 | 64.1 (8.5) | 63.3 | 57.7, 69.3 | 49.0-92.3 |
| Rwanda | 34 | 71.8 (8.5) | 73.0 | 64.3, 75.7 | 56.3-91.3 |

### 3.2. Results from adjusted linear regression models

Table 3 shows the adjusted associations between SBP and DBP and the concentrations of two metals, Pb and Cd. There was a statistically significant positive association between Pb exposure (log-transformed and not transformed) and SBP and DBB. The strongest association was observed as a 1-unit increase in log Pb value with an SBP increase by 2.36 mmHg (95% CI 0.51, 4.20). The adjusted associations between Cd exposure and SBP and DBP included the null value (Table 3). Overall, results suggested that exposure to Pb, but not Cd, was associated with increases in both systolic and diastolic BPs in the studied sample (Table 3).

**Table 3.** Primary results: Adjusted associations between outcomes of Systolic and Diastolic BP and exposures of Pb and Cd concentrations (K+ standardized)^1^.

| Outcomes | Metals exposures | N | Adjusted Estimate (mmHg) | 95% CI |
| --- | --- | --- | --- | --- |
| Systolic BP | Pb (natural log-transformed, K+ standardized) | 256 | 2.36 | 0.51, 4.20 |
| Systolic BP | Pb (untransformed, K+ standardized) | 256 | 1.03 | 0.93, 2.00 |
| Systolic BP | Cd (untransformed, K+ standardized) | 256 | -0.30 | -4.18, 3.58 |
| Diastolic BP | Pb (natural log-transformed, K+ standardized) | 256 | 1.42 | 0.16, 2.67 |
| Diastolic BP | Pb (untransformed, K+ standardized) | 256 | 0.61 | -0.02, 1.25 |
| Diastolic BP | Cd (untransformed, K+ standardized) | 256 | 0.59 | -2.04, 3.22 |

<sup>1</sup> Adjusted for: age, 5 household assets (color TV, radio, mobile phone, bank account, bicycle), food insecurity, education, diet diversity, secondhand smoke exposure, physical activity (METs/day), and BMI (kg/m<sup>2</sup>)

The sensitivity analysis (Table 4) presents the adjusted associations between BP (both systolic and diastolic) and the concentrations of Pb and Cd, without an additional adjustment for BMI. The association between Pb exposure (log-transformed and untransformed) and SBP remained statistically significant after removing BMI (e.g., 2.32 mmHg (95% CI 0.43, 4.21 for log-transformed Pb). After removing BMI, there was still no statistically significant association between Cd exposure and SBP. The association between Pb exposure (log-transformed) and DBP also remained statistically significant. For a unit increase in the natural log of Pb concentration, DBP increased by 1.42 mmHg (95% CI 0.15, 2.70). After removing BMI, there remained no statistically significant association between Cd exposure and DBP. The sensitivity analyses confirmed that the associations observed between Pb exposure and increased BP were robust, regardless of the inclusion or exclusion of BMI. Cd exposure continued to show no significant association with BP changes in the adjusted models.

**Table 4:** Sensitivity Analysis Results: Adjusted associations between outcomes of Systolic and Diastolic BP and exposures of Pb and Cd concentrations (K+ standardized), excluding BMI^1^.

| Outcomes | Metals exposures | N | Adjusted Estimate (mmHg) | 95% CI |
| --- | --- | --- | --- | --- |
| Systolic BP | Pb (natural log-transformed, K+ standardized) | 260 | 2.32 | 0.43, 4.21 |
| Systolic BP | Pb (untransformed, K+ standardized) | 260 | 0.99 | 0.03, 1.95 |
| Systolic BP | Cd (untransformed, K+ standardized) | 260 | 1.06 | -2.86, 4.98 |
| Diastolic BP | Pb (natural log-transformed, K+ standardized) | 260 | 1.42 | 0.15, 2.70 |
| Diastolic BP | Pb (untransformed, K+ standardized) | 260 | 0.60 | -0.05, 1.24 |
| Diastolic BP | Cd (untransformed, K+ standardized) | 260 | 1.43 | -1.20, 4.06 |

<sup>1</sup> Adjusted for: age, 5 household assets (color TV, radio, mobile phone, bank account, bicycle), food insecurity, education, diet diversity, secondhand smoke exposure, physical activity (METs/day)

Table 5 shows the effect modification results from the adjusted associations between outcomes of SBP and DBP and exposures of Pb and Cd concentrations. With interaction effects of exposure by age (median cutoff of <51 years and ≥51 years), and by research site (Guatemala, Peru, and Rwanda). Among women ≥51 years, there was an association with increased Pb per log-unit increase in Pb (3.41 mmHg (95% CI 0.79, 6.03). No other interaction models showed statistically significant interactions across age groups or countries, as all p-values for interaction were above 0.05 and 95% CIs included the null value.

**Table 5:** Effect modification Models. Adjusted associations between outcomes of Systolic and Diastolic BP and exposures of Pb and Cd concentrations (K+ standardized), interactions with age and research site.

| Personal exposures | N | Adjusted mean difference (mmHg) (95% CI) | p-value for interaction |
| --- | --- | --- | --- |
| **Log-transformed Pb with SBP** |  |  |  |
| Age <51 years | 130 | 1.15 (-1.56, 3.86) | 0.24 |
| Age ≥51 years | 130 | 3.41 (0.79, 6.03) |  |
| Guatemala | 109 | 3.18 (-0.24, 6.61) | 0.54 |
| Peru | 117 | 1.24 (-1.26, 3.75) |  |
| Rwanda | 34 | 3.72 (-1.53, 8.98) |  |

| **Log-transformed Pb with DBP** |  |  |  |
| --- | --- | --- | --- |
| Age < 51 years | 130 | 1.51 (-0.316, 3.34) | 0.89 |
| Age ≥51 years | 130 | 1.34 (-0.43, 3.11) |  |
| Guatemala | 109 | 1.33 (-0.96, 0.99) | 0.94 |
| Peru | 117 | 1.12 (-0.56, 2.79) |  |
| Rwanda | 34 | 1.78 (-1.73, 5.29) |  |
| **Untransformed Cd with SBP** |  |  |  |
| Age < 51 years | 130 | 1.74 (-3.97, 7.44) | 0.73 |
| Age ≥51 years | 130 | 0.36 (-5.06, 5.79) |  |
| Guatemala | 109 | 0.11 (-5.96, 6.19) | 0.68 |
| Peru | 117 | 1.86 (-3.81, 7.53) |  |
| Rwanda | 34 | 6.09 (-5.80, 17.98) |  |
| **Untransformed Cd with DBP** |  |  |  |
| Age < 51 years | 130 | 0.97 (-2.86, 4.79) | 0.76 |
| Age ≥51 years | 130 | 1.78 (-1.86, 5.41) |  |
| Guatemala | 109 | 1.00 (-3.03, 5.03) | 0.86 |
| Peru | 117 | 2.41 (-1.36, 6.17) |  |
| Rwanda | 34 | 2.61 (-5.28, 10.50) |  |

## 4. Discussion

Assessing metals’ exposure on SBP and DBP can provide insights into the broader implications of environmental health risks’ impacts on cardiovascular disease and inform public health strategies. A persistent public health concern in low- and middle-income countries is high BP, where cooking with solid biomass fuel is a prevalent practice. This produces dangerous levels of household air pollution, to which exposure can lead to significant morbidity and mortality, with air pollution accounting for nearly 1 in 6 deaths in children under 5 across Africa. Since women are most often the ones preparing meals on biomass cookstoves, they tend to have higher levels of exposure^19^.

These results provided an insightful overview of the baseline characteristics of the participants, highlighting important socioeconomic, health, and lifestyle factors. These factors are likely interrelated and may exacerbate disparities in health outcomes indicating significant health and socioeconomic challenges among the participants, such as overweight/obesity (65%), food insecurity with 36% experiencing mild and 17% moderate, and low educational attainment (77%). While most participants owned mobile phones (94%), fewer had access to other assets like radios (59%), televisions (49%), bicycles (31%), or bank accounts (30%). These findings reflect varying levels of socioeconomic status and suggest limited access to diverse communication tools and financial services in some groups.

These results also provided a comprehensive analysis of the associations between BP and exposure to Pb and Cd among adult women across Guatemala, Peru, and Rwanda, highlighting regional variations in BP levels among study participants, with Rwanda and Guatemala generally exhibiting higher SBP and DBP levels compared to Peru. In India, all participants that had valid blood pressure measurements at baseline and were largely normotensive (SBP <120 and DBP <80) (95%). The mean (SD) of SBP was 104.5 (9.1) mmHg and the mean (SD) of DBP was 61.5 (7.6) mmHg (Table 3). Distributions of baseline SBP and DBP by study sites are visualized in Figure 1 and 2. Thirteen pregnant women (2%) were considered as at hypertension stage 1, defined as SBP between 130 and 139 mmHg or DBP between 80 and 89 mmHg. Two individuals (<1% of the participants) were categorized as overly hypertensive, defined as SBP ≥140 mmHg or DBP ≥90 mmHg. The categorizations of normal blood pressure and of different stages of elevated blood pressure were not different between the two study sites^20^.

**Figure 1.**
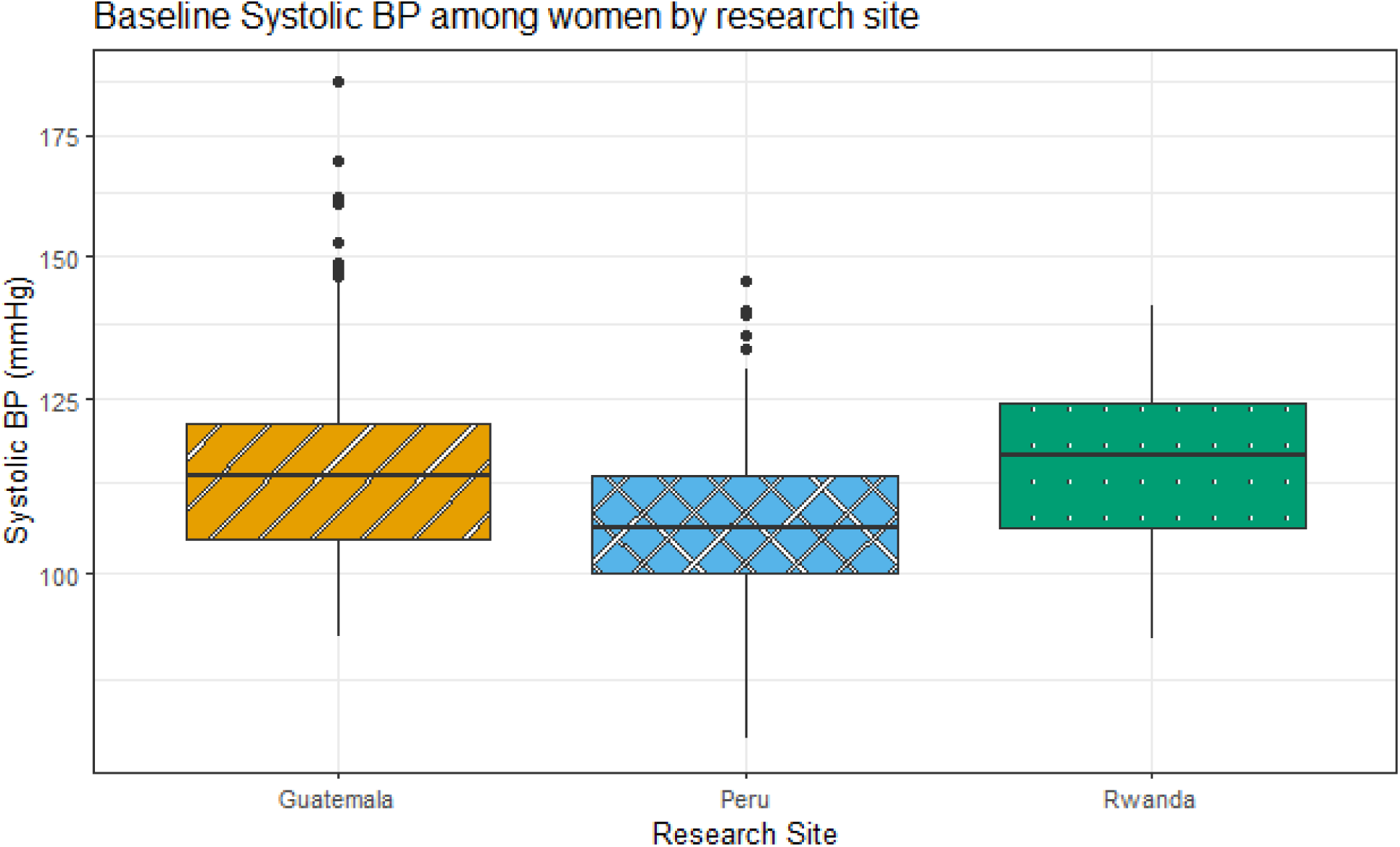
Boxplots of SBP (mmHg) by research site among other women with valid Pb and Cd measures at baseline (Guatemala, Peru, and Rwanda, N=272). In this boxplot Rwanda and Guatemala have the highest median values (116.3 mmHg and 113.3 mmHg) respectively compared to Peru which has 106 mmHg (values also shown in Table 2). The box represents the interquartile range with the line at the median value, the whiskers extend to data points within 1.5*IQR and the outliers are the points beyond the whiskers.

**Figure 2:**
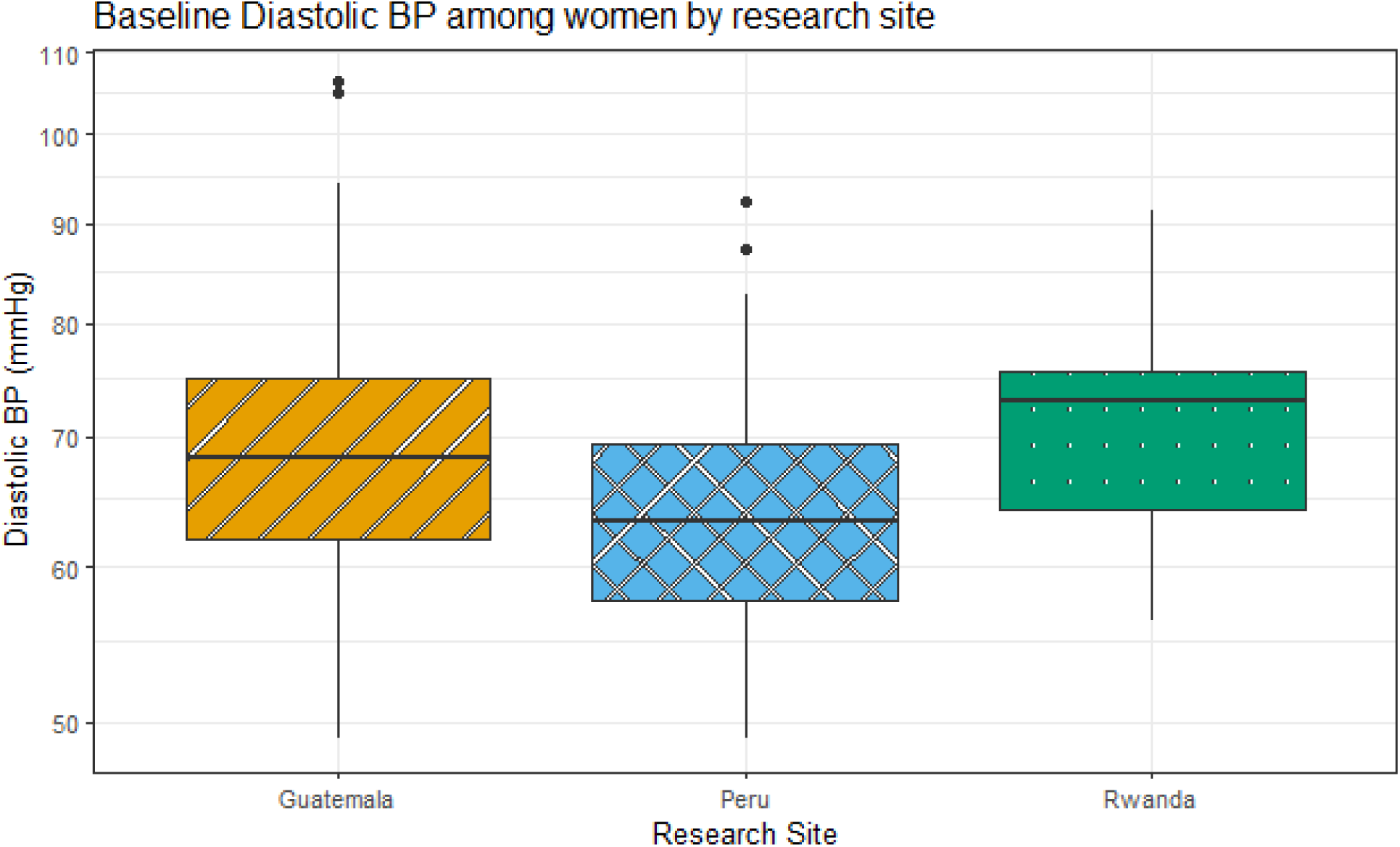
Boxplots of DBP (mmHg) by research site among other women with valid Pb and Cd measures at baseline (Guatemala, Peru, and Rwanda, N=272) which presents the median values of three countries/research sites. Rwanda had the highest median of 73mmHg followed by Guatemala and Peru with 68.3mmHg, 63.3mmHg respectively (values also shown in Table 2). The box represents the interquartile range with the line at the median value, the whiskers extend to data points within 1.5*IQR and the outliers are the points beyond the whiskers.

Pb exposure, as indicated by the median levels, showed regional disparities, with Guatemala having the highest median Pb levels, followed by Peru and Rwanda. Cd exposure, on the other hand, exhibited similar median levels across the three research sites, indicating consistent central tendencies. Pb exposure (log-transformed) showed statistically significant positive associations with both SBP and DBP, with as much as a 2.36 mmHg and 1.42 mmHg increases observed in SBP and DBP per supplemental table 1 respectively. Cd exposure, however, did not show significant associations with BP changes.

Sensitivity analyses, which excluded adjustment for BMI, confirmed the robustness of the associations between Pb exposure and BP changes. Even after excluding BMI, the positive associations remained statistically significant. Cd exposure continued to show no significant association with BP changes, even after removing BMI.

Current literature suggests that high BP is a persistent public health concern in low- and middle-income countries, where environmental factors such as air pollution from biomass fuel use significantly contribute to morbidity and mortality. Previous studies have demonstrated that household air pollution, which is common in low-and-middle income countries due to the use of solid biomass fuels for cooking, disproportionately affects women, who often spend more time near cookstoves. This exposure contributes to adverse health outcomes, including cardiovascular diseases. Air pollution alone accounts for nearly one in six deaths among children under five years across Africa^21, 22, 23^.

Prior research has also established a link between metal exposure and BP changes. For instance, a study in the United States that assessed associations of blood Pb, Cd, and mercury with resistant hypertension among adults in National Health and Nutrition Examination Survey, 1999–2018 showed that BP increased with higher concentrations of certain metals, including Pb (per 1μg/dL increase in blood Pb concentration, the proportion of resistant hypertension increased by 16% [adjusted odds ratio, 1.16; 95% confidence interval (CI) 1.01–1.32). This study adds to the existing body of knowledge by examining the associations between BP and metal exposure in diverse geographical regions, namely Guatemala, Peru, and Rwanda^24, 25^.Positive associations were also observed between exposure to metals and age, BMI and DBP in a study conducted in Peru^26^. In Korea a study found significant associations of blood Cd and Pb with higher hypertension prevalence and there was a significant interaction between blood Cd and physical activity on hypertension risk^27^.

These findings underscore the importance of understanding regional variations in BP levels and metal exposure for targeted public health interventions. The significant associations between Pb exposure and BP emphasize the need for policies and interventions aimed at reducing Pb exposure, especially in regions with high levels of exposure. Further research and monitoring are needed to better understand the mechanisms underlying the observed associations and to inform evidence-based interventions for preventing and managing hypertension and related cardiovascular diseases. Overall, these results contribute to our understanding of the complex relationships between environmental exposures, such as Pb and Cd, and cardiovascular health outcomes, highlighting the importance of multifaceted approaches to address public health challenges related to hypertension and metal exposure.

## 5. Conclusions

Adjusted associations between SBP and DBP and Pb concentrations showed statistically significant positive associations, even after adjusting for various factors, including BMI. The sensitivity analysis further confirmed the robustness of the observed associations between Pb exposure and increased BP, even after removing BMI. In summary, these findings highlight potential implications for public health interventions and further research into the determinants of BP variations across different geographical contexts. Additionally, the associations between Pb exposure and increased BP warrant attention and may have implications for environmental and occupational health policies.

## Acknowledgements

The HAPIN trial was funded by the U.S. National Institutes of Health (cooperative agreement 1UM1HL134590) in collaboration with the Bill & Melinda Gates Foundation [OPP1131279].

The investigators would like to thank the members of the advisory committee – Drs. Patrick Breysse, Donna Spiegelman, and Joel Kaufman - for their valuable insight and guidance throughout the implementation of the trial. We also wish to acknowledge all research staff and study participants for their dedication to and participation in this important trial.

A multidisciplinary, independent Data and Safety Monitoring Board (DSMB) appointed by the National Heart, Lung, and Blood Institute (NHLBI) monitored the quality of the data and protected the safety of patients enrolled in the HAPIN trial. The DSMB consisted of: Catherine Karr (Chair), Nancy R. Cook, Stephen Hecht, Joseph Millum, Nalini Sathiakumar (deceased), Paul K. Whelton, and Gail Weinmann and Thomas Croxton (Executive Secretaries). Program Coordination: Gail Rodgers, Bill & Melinda Gates Foundation; Claudia L. Thompson, National Institute of Environmental Health Sciences; Mark J. Parascandola, National Cancer Institute; Marion Koso-Thomas, Eunice Kennedy Shriver National Institute of Child Health and Human Development; Joshua P. Rosenthal, Fogarty International Center; Concepcion R. Nierras, NIH Office of Strategic Coordination – The Common Fund; Katherine Kavounis, Dong-Yun Kim, Barry S. Schmetter (deceased), and Antonello Punturieri, NHLBI.

This research represents the NIH’s contribution to the Global Alliance for Chronic Diseases (GACD) coordinated call for research on prevention and management of chronic lung diseases for 2016.

The findings and conclusions in this report are those of the authors and do not necessarily represent the official position of the US National Institutes of Health or Department of Health and Human Services.

## Statement of data sharing

Deidentified data associated with the paper will be deposited in Emory’s Dataverse data repository. A DOI for the data will be created, to allow citation with publication of the paper. Dataverse is widely accessible and provides long-term access to the public and to related research communities.

